# CT Coronary Angiography Identifies a Shear-Stress Signature of Spontaneous Coronary Artery Dissection: A Case–Control Study

**DOI:** 10.64898/2026.08.12.26360329

**Authors:** Mingzi Zhang, Lucy McGrath-Cadell, Stephanie Hesselson, Chi Shen, Ramtin Gharleghi, Nicholas Collins, David WM Muller, Jason C. Kovacic, Robert M Graham, Susann Beier

## Abstract

**Background:** Spontaneous coronary artery dissection (SCAD) causes acute coronary syndrome that predominantly affects women. It is not known why SCAD occurs in specific coronary artery segments. We aimed to identify anatomical and hemodynamic factors that lead to SCAD.

**Methods:** We studied 36 women with angiographically-confirmed SCAD from more than 20 hospital sites and 75 sex- and ethnicity-matched control participants with normal coronary anatomy. Coronary arteries were reconstructed from computed tomography coronary angiography (CTCA) to quantify vessel geometry (curvature, diameter, torsion) and flow-derived metrics (time-averaged endothelial shear stress [TAESS], topological shear variation index [TSVI], oscillatory shear index [OSI], and relative residence time [RRT]) at the tree (left/right), territory (LAD, LCx, RCA), and lesion levels.

**Results:** Compared with controls, SCAD-affected coronary arteries had greater curvature and higher TAESS and TSVI at the whole-tree level (all p≤0.007). At the vessel (territory) level, SCAD-affected arteries were smaller in average diameter and showed higher curvature, TAESS, and TSVI than matched control vessels (all p≤0.047). Within the same patient, SCAD lesion segments were characterized by smaller diameter, lower torsion, and higher TAESS and TSVI than non-affected segments from the same coronary tree (all p≤0.001; curvature borderline). A model combining curvature, TAESS, and TSVI discriminated SCAD from controls with AUC 0.95 (left tree) and 0.97 (right tree); adding diameter yielded AUCs >0.91 at the territory level.

**Conclusions:** SCAD was associated with a reproducible multi-scale signature of smaller vessel caliber and higher, more variable endothelial shear stress supporting a hemodynamic contribution to SCAD clustering in specific coronary arteries and segments.

**Graphical abstract:** 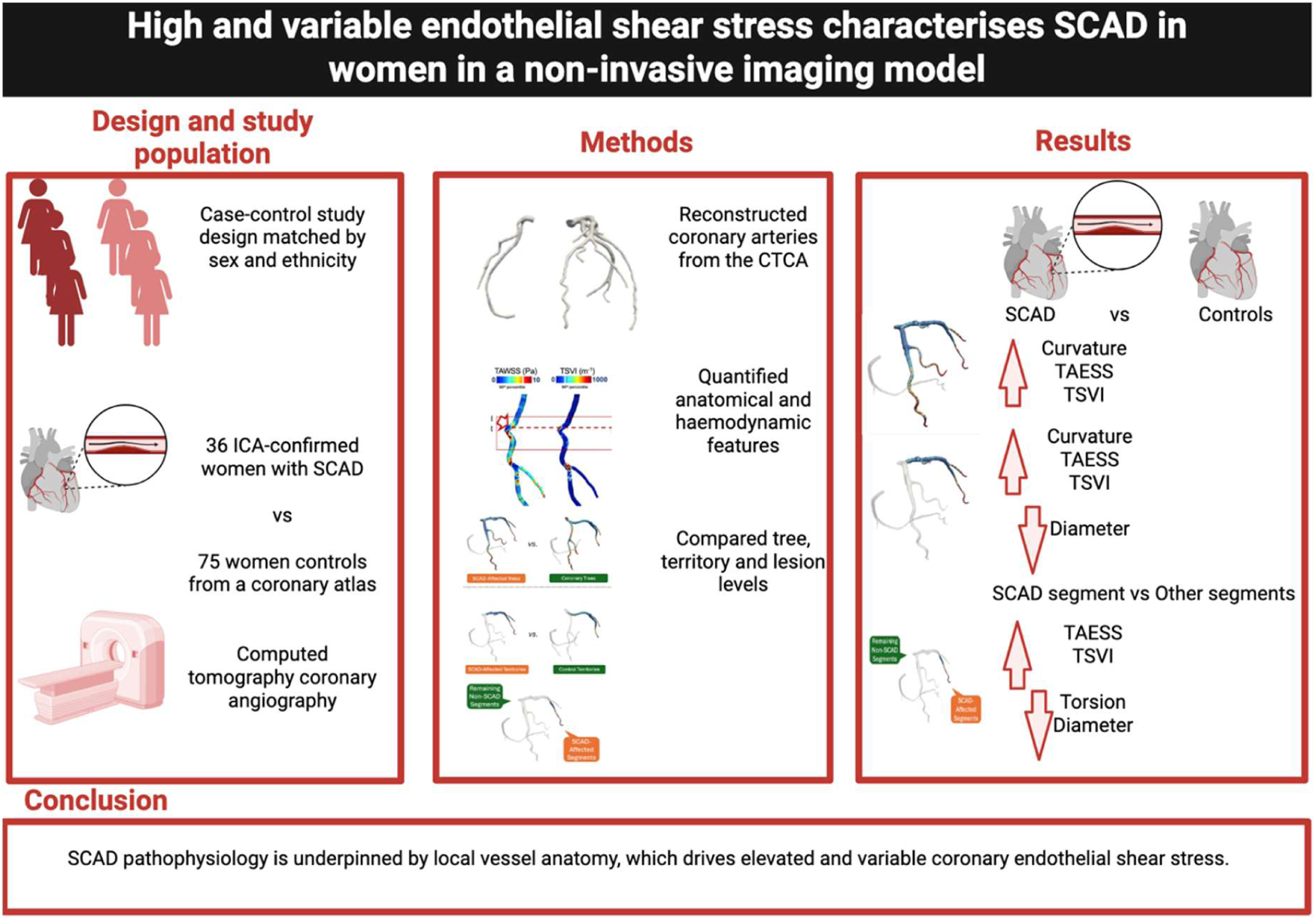

## Introduction

Spontaneous coronary artery dissection (SCAD) accounts for up to 35% of acute coronary syndromes (ACS) in women under 50 years. More than 90% of cases are women, who are on average 20-30 years younger (average age 45-52 years) than patients with their first presentation of atherosclerotic ACS.^1^ SCAD is most commonly due to the spontaneous development of a hematoma within the media of a coronary artery (angiographic type 2 and 3 SCAD, ∼70% prevalence) and may also involve intimal disruption (type 1 SCAD, ∼30% prevalence).^2^ For survivors, SCAD heals spontaneously, generally within 30 days.^3^ Genetics studies suggest a contribution from coronary vessel extracellular matrix and collagen vulnerabilities that enhance the propensity for coronary vasa vasorum to rupture and bleed.^4^ However, it remains unclear why, in genetically vulnerable individuals, SCAD occurs at certain locations within the coronary artery tree.

Computational fluid dynamics (CFD) methods are used to assess endothelial shear stress caused by hemodynamic forces. This involves generating three-dimensional models of coronary arteries from imaging data and then calculating blood flow within them.^5^ Time-averaged endothelial (or wall) shear stress (TAESS) is a validated metric, abnormalities of which are associated with a higher risk of future arterial diseases (e.g. aneurysm, atherosclerosis).^5–7^

A recent exploratory study proposed that, based on CFD-determined parameters, greater curvature, torsion and TAESS profiles, which are associated with increased local blood flow disturbances, play a pathophysiological role in SCAD.^8^ In fact, the emerging concept of acute Mechano–Cardiac Coronary Artery Disruption (AMCAD) has been hypothesized as a unifying entity across SCAD and traumatic arterial dissections, highlighting the risk of cervical artery dissections linked to excessive neck movements.^9^ It was even suggested that the term “spontaneous” should be reconsidered in some cases.^9^

Local anatomical characteristics, such as curvature and torsion, govern the resulting blood flow dynamics marked by metrics such as ESS. In fact, tortuosity has been repeatedly highlighted as a hallmark of SCAD cases. However, it has previously been demonstrated that tortuosity, as predominantly measured in clinical observations, introduces significant errors when aiming to capture the complex, twisted nature of coronary curvature in three-dimensional space.^10^ Instead, it was demonstrated that the average curvature is a more robust three-dimensional measure and should be applied.

As SCAD predominantly affects women, a fundamental need for sex-specific considerations has been demonstrated, driven by well-documented anatomical differences in male and female patients^11^, which in turn alter relevant blood flow patterns in women compared to men.^12–14^.

Therefore, there is a great opportunity in the field to better understand mechanical forces within the coronaries, and to overcome persistent literature shortfalls to date, including sex-specific studies, single-center studies, or investigations lacking matched control groups for meaningful and adequate sample sizes for statistical confidence.

We therefore performed a matched case–control analysis in females with confirmed SCAD, comparing anatomical and hemodynamic parameters across the entire coronary tree, individual territories, and discrete lesion segments. We show that SCAD pathophysiology is marked at the coronary tree, territory, and segment levels by distinct anatomical and hemodynamic characteristics compared to non-SCAD patients and vessels, with higher and more variable endothelial shear stress evident across all SCAD-affected domains.

## Methods

### Study population

The subjects were a subset of the VCCRI Arteriopathies and SCAD Cohort (VASC) collected from more than 20 hospital sites and many different imaging centers, including, in particular, St Vincent’s Hospital, Sydney and John Hunter Hospital, Newcastle. Detailed VASC recruitment and data collection are outlined in **Supplement 1**. The study was approved by the St Vincent’s Hospital Human Research Ethics Committee (2019/ETH03171). VASC cases that had a CTCA post-SCAD were identified and included in this study. All cases provided written informed consent. We limited our study group to women only. Participants were predominantly Caucasian (33 cases; 92%), with 3 cases (8%) being Asian. Normal control subjects were selected from the Coronary Atlas dataset^11^, previously established for characterizing 3D coronary geometry and blood flow in patients with suspected CAD and zero calcium scores. Use of this dataset was approved by the institutional ethics committees of the University of New South Wales (Ref. HC190145) and the University of Auckland (Ref. 02296). Only subjects without CAD and with a zero-calcium score and no abnormal CTCA findings were included.

### CTCA and ICA imaging

CTCA studies were performed according to the local imaging protocols at the respective imaging departments, as detailed in **Supplement 2**. CTCA studies were manually checked for sufficient image quality for subsequent vessel segmentation and reconstruction prior to inclusion in the study, this included only including CTCAs with a slice thickness of ≤1mm and excluding CTCA with artefacts (**Figure 1**). The studies were performed at varying times after the SCAD event, with the majority performed outside the acute SCAD period (>30 days).

**Figure 1.**
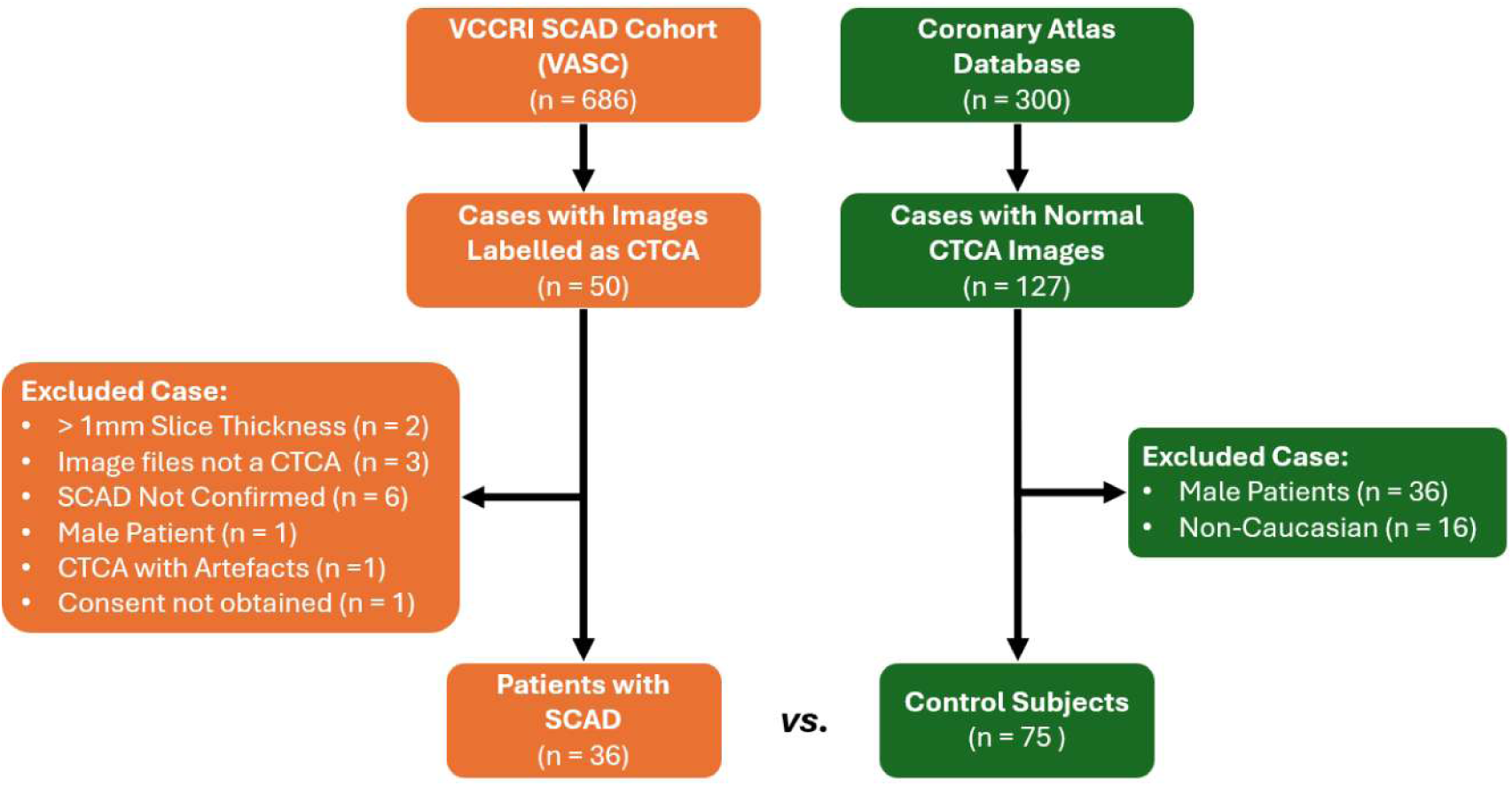
Patient inclusion and exclusion diagram.

The original diagnostic invasive coronary angiogram (ICA) from the time of the index SCAD event was obtained for all cases. The ICA diagnosis of SCAD was confirmed by the initial interventionalist and independently verified through our core laboratory review of the images.

Each included case had been diagnosed as SCAD by the initial interventional cardiologist who performed the diagnostic ICA. The director of the cardiac catheter laboratory at St Vincent’s Hospital, an experienced (>30 years) senior interventional cardiologist with extensive experience in diagnosing and managing SCAD cases (D.W.M.M.) and a cardiologist specializing in SCAD with more than 8 years’ experience (L.M-C.), jointly reviewed all the original ICA images and confirmed the dissection. Cases that could not be confirmed on ICA or intravascular imaging were excluded from the analysis (**Figure 1**). We utilized modified ICA diagnostic criteria for SCAD based upon that described by Saw et al.^2^ with type 1 SCAD involving intimal disruption with contrast staining of the vessel wall, type 2 SCAD involving a long segment of luminal stenosis secondary to intramural hematoma, type 3 SCAD being a discrete luminal narrowing, also due to an intramural hematoma. Type 4 SCADs, an additional criterion that was recognized later, representing complete artery occlusion, were also evaluated. The artery, segment and location of the dissection were determined, as well as the length, type of SCAD (1-4) and TIMI flow through the affected vessel. This enabled direct correlation between the exact location of the SCAD and the CFD at that location compared with the rest of the vessel and the coronary tree.

Cases with atherosclerotic plaque on invasive angiogram were excluded from the analysis. One case had been confirmed on intravascular ultrasound during the initial angiogram, despite the concurrent presence of atheroma remote from the SCAD location, so was retained in the analysis.

### Coronary anatomy and hemodynamic characterization

Detailed computational settings and definitions of anatomic and hemodynamic metrics are presented in **Supplement 3 and 4**. Briefly, coronary arteries were auto-segmented from CTCA using a previously benchmarked deep-learning pipeline^15^, followed by expert verification. The digital reconstructions enabled automated extraction of vessel-specific anatomic characteristics, including vessel diameter, absolute curvature (as the best measure of tortuosity)^10^, and torsion,^16,17^ based on the Vascular Modelling ToolKit (v1.4).

We evaluated four complementary SCAD-related hemodynamic metrics reported in the literature using computational blood flow simulations following standard methods previously described ^18–20^, including TAESS, OSI, RRT, and TSVI.^8^ TAESS is a key parameter associated with endothelial cell dysfunction if outside of standard physiological ranges, calculated as the shear stress magnitude over a cardiac cycle. OSI and RRT describe multi-directional fluctuations and low and oscillatory shear stress on the endothelium over a cardiac cycle, both of which are implicated in atherosclerosis and aneurysm progression.^6,7^ TSVI quantifies variability in the contraction and expansion forces exerted by shear stress on endothelial cells during a cardiac cycle, a surrogate risk indicator of adverse clinical events.^8,21^

### Statistical analysis

Statistical analyses were performed using Python (v3.12) with the following packages: SciPy (v1.16) for statistical functions, scikit-learn (v1.7) for bootstrap resampling, and matplotlib (v3.10) for visualization. Statistical analyses used are detailed in the **Supplement 5.**

## Results

### Patient characteristics

Thirty-six patients with SCAD (41 lesions; median age 46 years) and 75 controls (median age 58 years) met the selection criteria and were included in the final analysis (**Figure 1 and Table 1**). Among the cohort, three cases were identified as being of Asian ethnicity. The results remained unchanged when these cases were excluded; therefore, they were included in the final analyses. The only baseline characteristic that differed significantly between the two groups was smoking status (p=0.047, **Table 1**). Other relevant comorbidities and characteristics in the SCAD group are detailed in **Table 1**, and angiographic SCAD details are as in **Table 2**. The timing between SCAD presentation and CTCA imaging varied widely (7-390 days) with a median follow-up time of 72 days.

**Table 1.**
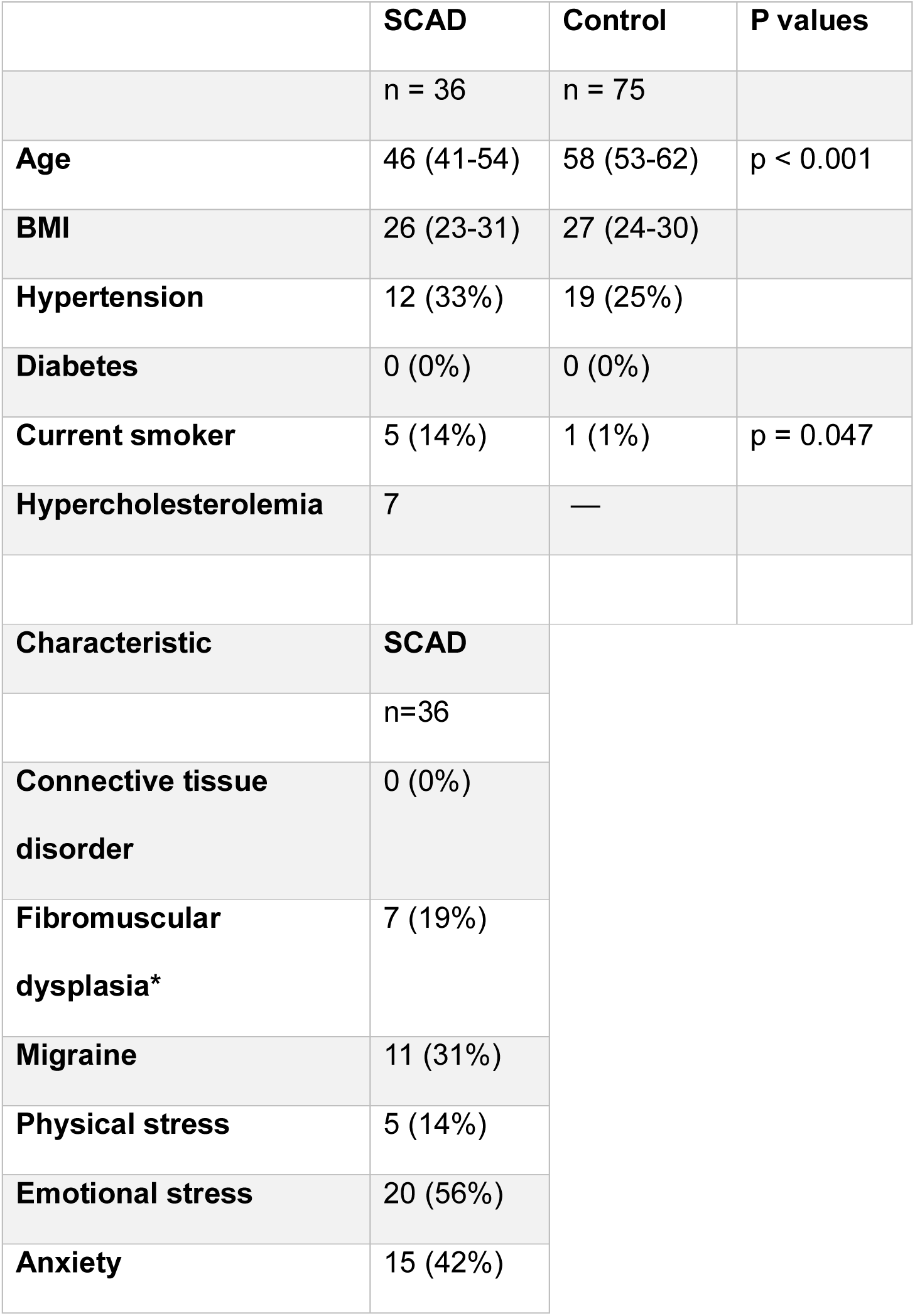
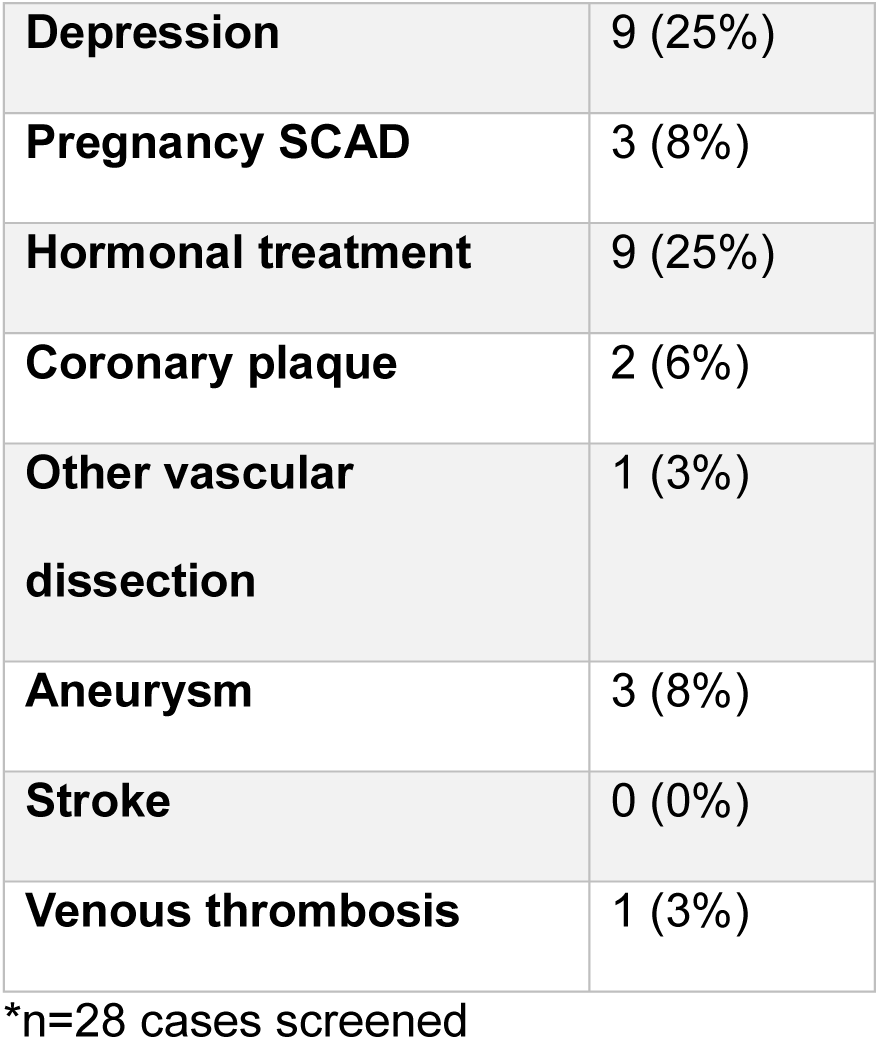
Baseline characteristics comparison between SCAD patients and controls and additional comorbidities in SCAD patients.

|  | SCAD | Control | P values |
| --- | --- | --- | --- |
|  | n = 36 | n = 75 |  |
| **Age** | 46 (41-54) | 58 (53-62) | p < 0.001 |
| **BMI** | 26 (23-31) | 27 (24-30) |  |
| **Hypertension** | 12 (33%) | 19 (25%) |  |
| **Diabetes** | 0 (0%) | 0 (0%) |  |
| **Current smoker** | 5 (14%) | 1 (1%) | p = 0.047 |
| **Hypercholesterolemia** | 7 | — |  |
| **Characteristic** | **SCAD** |  |  |
|  | n=36 |  |  |
| **Connective tissue disorder** | 0 (0%) |  |  |
| **Fibromuscular dysplasia\*** | 7 (19%) |  |  |
| **Migraine** | 11 (31%) |  |  |
| **Physical stress** | 5 (14%) |  |  |
| **Emotional stress** | 20 (56%) |  |  |
| **Anxiety** | 15 (42%) |  |  |

|  |  |
| --- | --- |
| **Depression** | 9 (25%) |
| **Pregnancy SCAD** | 3 (8%) |
| **Hormonal treatment** | 9 (25%) |
| **Coronary plaque** | 2 (6%) |
| **Other vascular**<br>**dissection** | 1 (3%) |
| **Aneurysm** | 3 (8%) |
| **Stroke** | 0 (0%) |
| **Venous thrombosis** | 1 (3%) |

\*n=28 cases screened

**Table 2.**
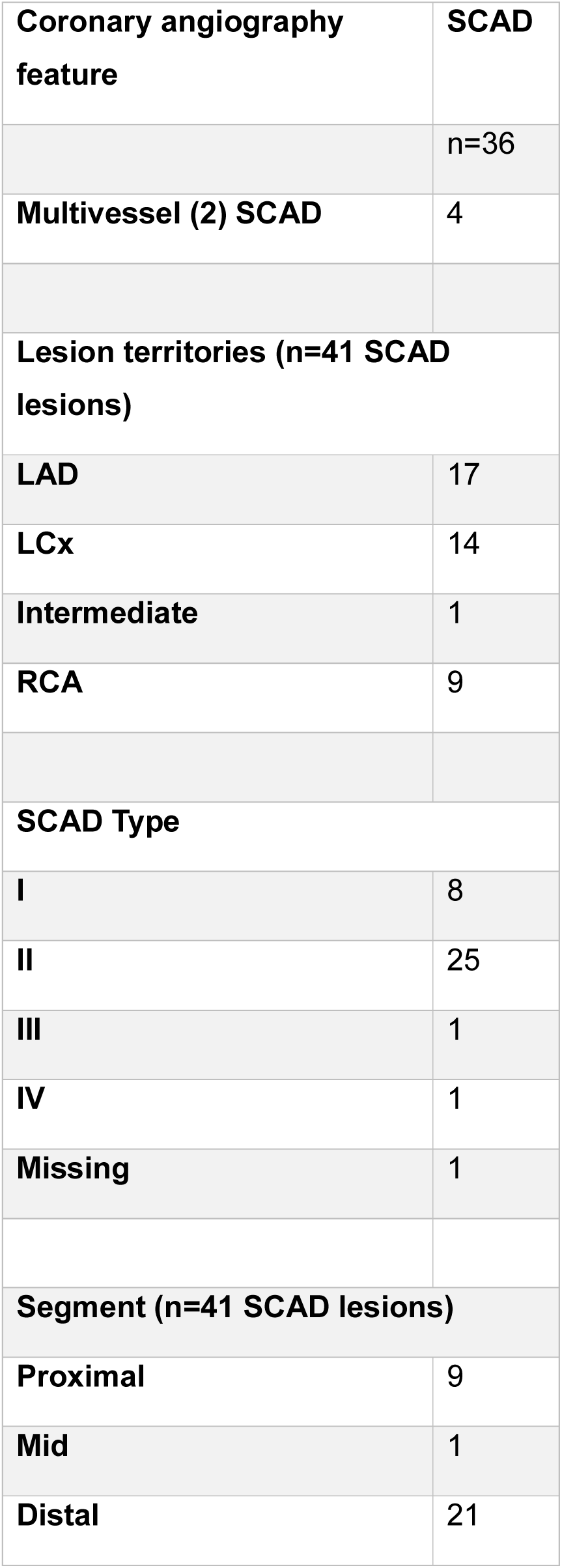
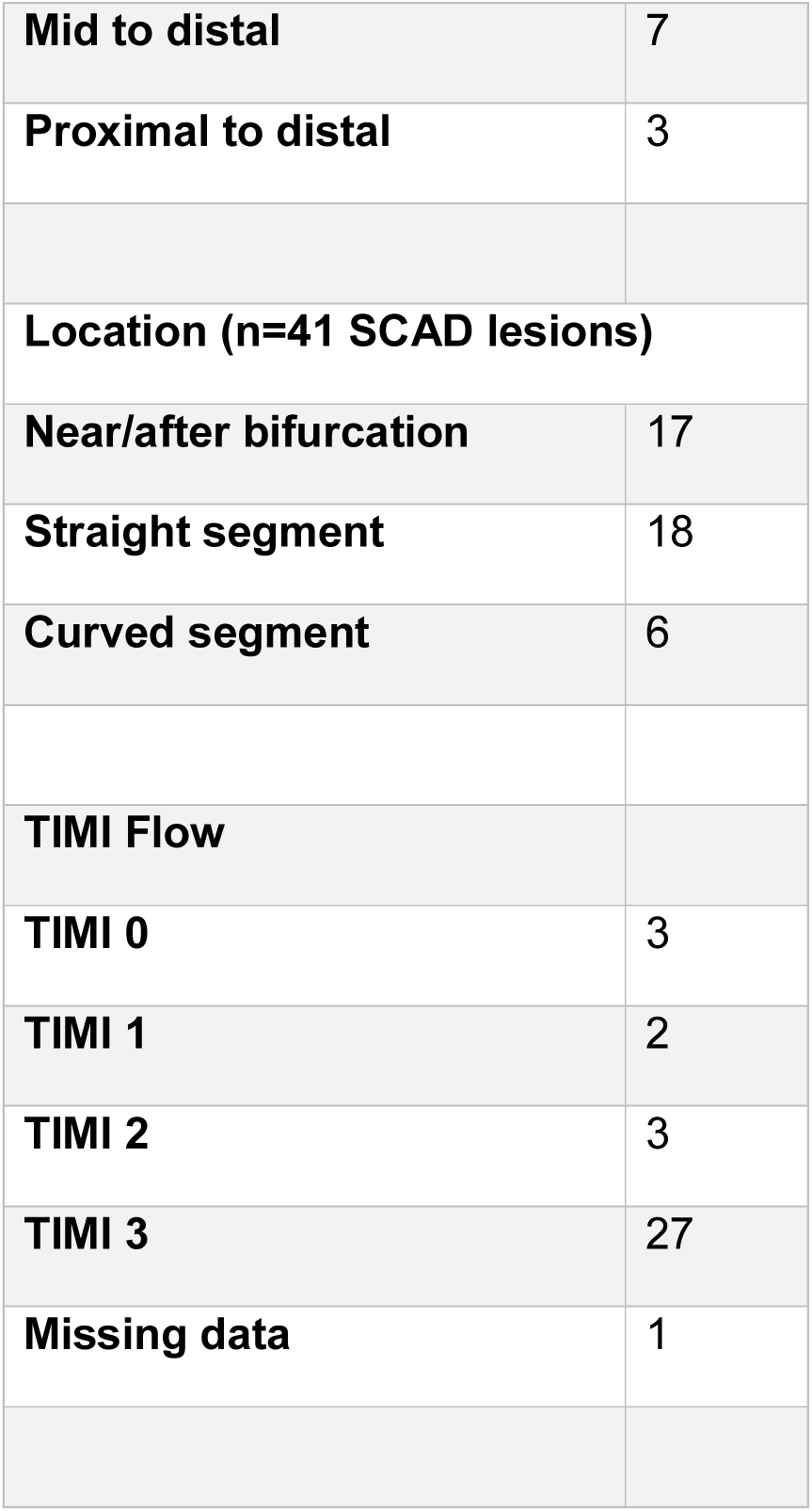
Invasive coronary angiography features in SCAD patients.

| Coronary angiography feature | SCAD |
| --- | --- |
|  | n=36 |
| Multivessel (2) SCAD | 4 |
| Lesion territories (n=41 SCAD lesions) |  |
| LAD | 17 |
| LCx | 14 |
| Intermediate | 1 |
| RCA | 9 |
| SCAD Type |  |
| I | 8 |
| II | 25 |
| III | 1 |
| IV | 1 |
| Missing | 1 |
| Segment (n=41 SCAD lesions) |  |
| Proximal | 9 |
| Mid | 1 |
| Distal | 21 |
| **Mid to distal** | 7 |
| **Proximal to distal** | 3 |
| **Location (n=41 SCAD lesions)** |  |
| **Near/after bifurcation** | 17 |
| **Straight segment** | 18 |
| **Curved segment** | 6 |
| **TIMI Flow** |  |
| **TIMI 0** | 3 |
| **TIMI 1** | 2 |
| **TIMI 2** | 3 |
| **TIMI 3** | 27 |
| **Missing data** | 1 |

Interestingly, no statistically significant differences in any metrics between acute SCAD (CTCA within 30 days of ICA diagnosis of SCAD), and healed SCAD (CTCA outside 30 days of ICA diagnosis of SCAD) (**Supplement 6, Table S1).** Thus, despite the variability in time of CTCA acquisition relative to SCAD event, this has no impact on the overall findings, which remain consistent between the acute SCAD group (≤30 days between SCAD and CTCA) and the healed SCAD group (>30 days between SCAD and CTCA). We also observed no significant differences in any metrics between type I SCAD (angiographic double lumen) and other SCAD types (intramural hematoma) (**Supplement 7, Table S2**).

### SCAD-affected coronary trees are marked by greater curvature with high TAESS and TSVI

Consistent with the territory-based analysis, the SCAD-affected left coronary artery trees showed significantly greater tree-averaged curvature than those of the control group: 0.45 ± 0.25 vs. 0.19 ± 0.02 x 10^3^ mm^−1^ (p<0.001), with a large effect size *r_rb_* = 0.91, 95% confidence interval (CI): [0.81, 0.98]. Similarly, significantly higher tree-averaged TAESS (*r_rb_* = 0.58, 95% CI: [0.36, 0.79]) and TSVI (*r_rb_* = 0.48, 95% CI: [0.20, 0.75]) in the SCAD than the control group (p<0.001 for both parameters) remained persistent.

The right coronary tree in the SCAD group also exhibited significantly greater curvature (p<0.001), TAESS (p=0.007), and TSVI (p<0.001), while no significant differences were observed in diameter (p=0.39), torsion (p=0.707), and RRT (p=0.407). Full details and visual comparisons are provided in **Table 3a** and **Figure 2 (A)**.

**Figure 2.**
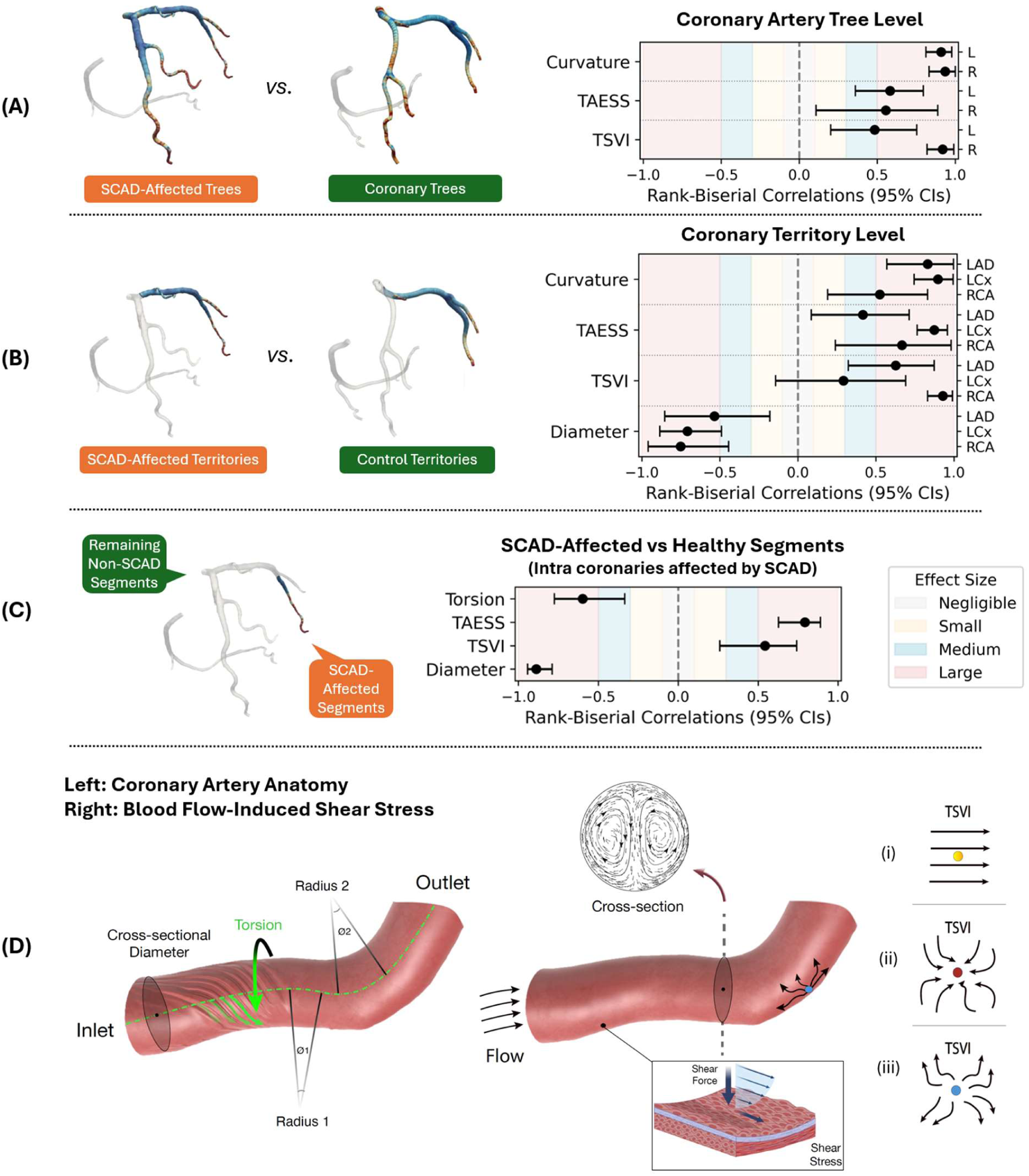
(A): Coronary-tree level comparison (Left L vs Right R) suggests that curvature and time-averaged endothelial shear stress (TAESS) have a large effect size, and topological shear variation index (TSVI) a medium-to-large effect size between the SCAD-affected and control coronary trees. (B): Territory-level comparison suggests coronary artery diameters are significantly smaller in SCAD-affected territories, in addition to the greater curvature, TAESS, and TSVI. (C): Within the SCAD-affected coronary trees, the SCAD-affected segments no longer differed from the non-SCAD-affected segments in curvature, but still in diameter, TAESS, TSVI, and the average torsion. (D): Schematic illustration of coronary artery anatomy (left) and flow-derived shear metrics (right). Left: centerline-based geometric descriptors, including cross-sectional diameter and local vessel curvature defined by the inner and outer radii of curvature (Radius 1 and Radius 2), as a reliable measure of vascular tortuosity, as well as torsion describing axial vessel rotation along its centerline. Right: blood flow-induced ESS patterns and the cross-sectional secondary flow profile, together with conceptual basis of the TSVI. Under predominantly unidirectional laminar flow (i) TSVI is low due to aligned shear vectors. In disturbed or recirculating flow conditions, shear vectors vary markedly in directions, resulting in elevated TSVI, associated with shear stress-induced endothelial contraction (ii) or expansion (iii).

**Table 3a.** Tree- and territory-level differences in curvature, diameter, torsion, time-averaged endothelial shear stress (TAESS), relative residence time (RRT), and topological shear variation index (TSVI) between SCAD (n=36) and control patients (n=75).

| Metrics | Groups | Tree-Level |  | Territory-Level |  |  |
| --- | --- | --- | --- | --- | --- | --- |
|  |  | Left | Right | LAD | LCx | RCA |
| **Curvature**<br>**(10<sup>3</sup> mm<sup>-1</sup>)** | SCAD | 0.450 (0.245) | 0.442 (0.274) | 0.507 (0.279) | 0.403 (0.192) | 0.454 (0.328) |
|  | Control | 0.193 (0.02) | 0.173 (0.02) | 0.189 (0.029) | 0.183 (0.032) | 0.228 (0.04) |
|  | P values | **<0.001\*** | **<0.001\*** | **<0.001\*** | **<0.001\*** | **<0.001\*** |
| **Diameter**<br>**(mm)** | SCAD | 2.450 (0.407) | 2.678 (0.476) | 2.004 (0.943) | 2.004 (0.313) | 1.547 (0.289) |
|  | Control | 2.415 (0.256) | 2.673 (0.301) | 2.417 (0.406) | 2.54 (0.408) | 2.13 (0.436) |
|  | P values | 0.500 | 0.398 | **<0.001\*** | **<0.001\*** | **<0.001\*** |
| **Torsion**<br>**(mm<sup>-1</sup>)** | SCAD | 79.834 (97.687) | 106.867<br>(149.049) | 59.021 (48.211) | 44.271 (33.268) | 204.157<br>(294.386) |
|  | Control | 23.098 (7.777) | 32.704 (8.749) | 23.474 (7.201) | 20.503 (4.52) | 33.133 (13.541) |
|  | P values | 0.384 | 0.707 | **0.009\*** | **<0.001\*** | 0.056 |
| **TAESS**<br>**(Pa)** | SCAD | 1.803 (1.195) | 2.176 (1.516) | 3.133 (3.882) | 3.327 (2.307) | 5.111 (2.847) |
|  | Control | 0.987 (0.371) | 1.232 (0.4) | 1.013 (0.415) | 0.949 (0.516) | 1.925 (1.216) |
|  | P values | **<0.001\*** | **0.007\*** | **0.007\*** | **<0.001\*** | **0.002\*** |
| **RRT**<br>**(Pa<sup>-1</sup>)** | SCAD | 1.839 (1.325) | 1.768 (1.637) | 1.615 (1.428) | 0.896 (0.512) | 0.969 (1.17) |
|  | Control | 1.662 (0.651) | 1.335 (0.946) | 1.603 (0.755) | 1.723 (0.653) | 1.18 (1.169) |
|  | P values | 0.890 | 0.407 | 0.411 | **<0.001\*** | 0.167 |
| **TSVI**<br>**(mm<sup>-1</sup>)** | SCAD | 0.166 (0.065) | 0.121 (0.060) | 0.229 (0.117) | 0.175 (0.101) | 0.197 (0.136) |
|  | Control | 0.129 (0.028) | 0.037 (0.023) | 0.128 (0.03) | 0.114 (0.028) | 0.051 (0.039) |
|  | P values | **<0.001\*** | **<0.001\*** | **<0.001\*** | **0.047\*** | **<0.001\*** |

Note: Values in the parentheses are the standard deviations of the respective metrics. \* Indicates statistical significance.

Using the three identified features, curvature, TAESS, and TSVI, to identify previous SCAD episodes, yielded receiver operating characteristics (ROC) with high areas under the curve (AUCs) values for both the left (0.95, 95% CI: [0.90, 0.99]) and right (0.97, 95% CI: [0.93, 0.99]) coronary artery tree (**Figure 3**, upper panels). Given the small sample size and lack of external validation, these findings are considered exploratory.

**Figure 3.**
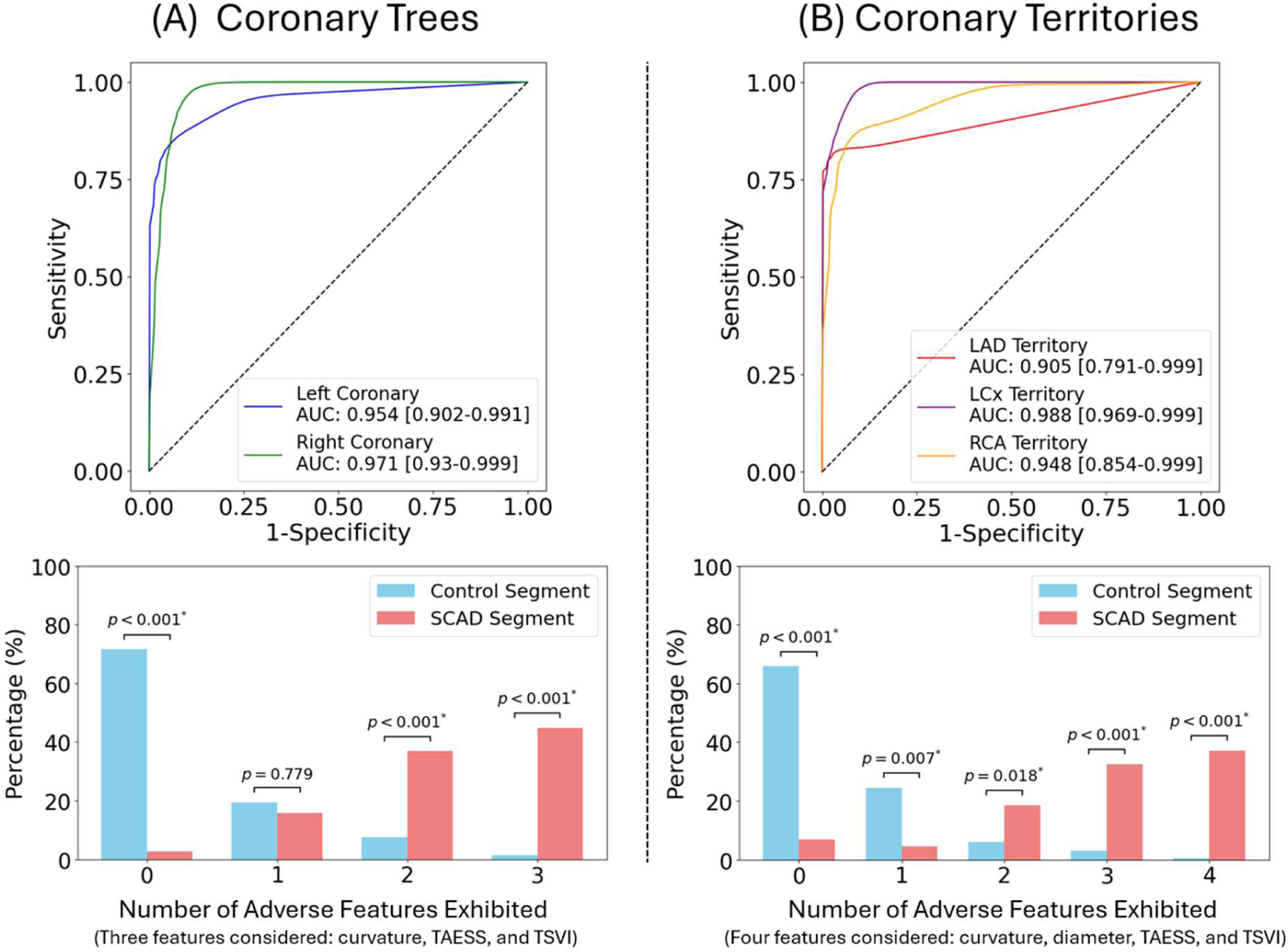
Receiver operating characteristics (ROC) curves of employing multiple features with medium to large effect sizes to predict SCAD occurring in the left or right coronary artery trees (A) and the left anterior descending (LAD), left circumflex (LCx), and right coronary artery (RCA) territories (Top). The proportion of normal and SCAD-affected segments by number of adverse features exhibited (Bottom). At the coronary tree level, three adverse features considered were (1) curvature, (2) time-averaged endothelial shear stress (TAESS), and (3) topological shear variation index (TSVI); and at the territory level, four features considered were features (1), (2), (3), and (4) diameter. AUC = area under the ROC curve; CI = confidence intervals.

The number of adverse features was higher in SCAD patients than in controls: 107 (71.5%) of the controls exhibited no single adverse feature, compared with only 2 (2.6%) of the SCAD patients (p<0.001). Additionally, only 2 (1.4%) of the controls exhibited all three adverse features, compared with 32 (44.7%) of the SCAD patients (p<0.001; **Figure 3**, lower panels). The optimal cut-offs along with their respective sensitivities and specificities for each feature are presented in **Supplement 8, Table S3**.

Although OSI differed statistically between the SCAD and control groups (p=0.037), the mean values (e.g., 0.014 ± 0.01 for the left coronary artery tree, Table 2a) were >30 times lower than the upper bound of OSI (<0.5). Thus, OSI was not considered in the subsequent diagnostic performance analyses. See **Supplement 9, Figure S2** for the complete set of effect sizes for all metrics.

### SCAD-affected coronary territories are marked by small diameter and greater curvature with high TAESS and TSVI

The territory-level comparisons revealed that, in addition to greater curvature (p<0.001), TAESS (p<0.007), and TSVI (p<0.047), the entire SCAD-affected vessels, including the segments not affected by the SCAD, had significantly smaller average diameters (in mm) compared to control territories (LAD: 2.00 ± 0.94 vs. 2.42 ± 0.41; LCx: 2.00 ± 0.31 vs. 2.54 ± 0.41; RCA: 1.55 ± 0.29 vs. 2.13 ± 0.44; all p<0.001, **Table 3b** and **Figure 2 (B)**).

**Table 3b.** Intra-patient differences in curvature, diameter, torsion, time-averaged endothelial shear stress (TAESS), relative residence time (RRT), and topological shear variation index (TSVI) between SCAD-affected (n=41 lesions) and the remaining normal segments.

| Metrics | Lesion Segments | Normal Segments | Effect Sizes (95% CIs) | P Values |
| --- | --- | --- | --- | --- |
| Curvature ($10^3 \text{ mm}^{-1}$) | 0.478 (0.262) | 0.442 (0.245) | 0.338 (0.009, 0.601) | 0.053 |
| Diameter (mm) | 1.722 (0.686) | 2.649 (0.451) | -0.888 (-0.942, -0.789) | <0.001* |
| Torsion ($\text{mm}^{-1}$) | 83.91 (147.668) | 101.342 (92.93) | -0.598 (-0.775, -0.334) | <0.001* |
| TAESS (Pa) | 3.619 (3.187) | 1.493 (1.006) | 0.793 (0.627, 0.89) | <0.001* |
| RRT ($\text{Pa}^{-1}$) | 1.212 (1.14) | 1.941 (1.463) | -0.708 (-0.842, -0.493) | <0.001* |
| TSVI ($\text{mm}^{-1}$) | 0.202 (0.116) | 0.152 (0.061) | 0.543 (0.26, 0.741) | 0.001* |

Note: Values in the parentheses are the standard deviations of the respective metrics unless otherwise noted. Effect sizes are reported using the matched-rank biserial correlation test. \* Indicates statistical significance.

While vessel-averaged torsion was significantly higher in the SCAD-affected LAD (p<0.01) and LCx (p<0.001) territories, the difference in the RCA (p=0.056) was borderline significant compared to its respective control.

Thus, only diameter, curvature, TAESS, and TSVI were included in the diagnostic performance analyses. Based on optimal cut-offs derived for the four features (see **Supplement 8, Table S3**), multi-variable logistic regression exploratory analysis demonstrated highly favorable classification performance for the SCAD-affected territories, including LCx (AUC: 0.99, 95% CI: [0.97, 0.99]), RCA (AUC: 0.95, 95% CI: [0.85, 0.99]) and LAD (AUC: 0.91, 95% CI: [0.79, 0.99]).

Interestingly, 65.8% of control vessels had not a single adverse feature compared to only 7.0% of SCAD-affected vessels (p<0.001). In contrast, 0.5% of the control vessels and 37.2% of the SCAD-affected vessels exhibited all four adverse features (p<0.001, **Figure 3**, lower panels).

### SCAD-affected vessel segments marked by small diameter and large torsion with high TAESS and TSVI

SCAD-affected segments exhibited a statistically smaller diameter (p<0.001, *r_rb_* = −0.88, 95% CI: [−0.94, −0.79]), critically higher TAESS (p<0.001, *r_rb_* = 0.79, 95% CI: [0.63, 0.89]), and higher TSVI (p=0.001) compared to non-affected segments. Similarly, the segment-averaged torsion within the SCAD-affected regions (83.91 ± 147.67 mm^−1^) was significantly lower than that in the remaining regions (101.34 ± 92.93 mm^−1^, p<0.001), with a large effect size *r_rb_* = −0.59, 95% CI: [−0.77, −0.33] (**Table 3b** and **Figure 2 (C)**).

Curvature was greater in the SCAD-affected segments (0.48 ± 0.26 10^3^ mm^−1^) compared to the remaining segments (0.44 ± 0.25 10^3^ mm^−1^) within the same coronary trees, although of borderline statistical significance (p=0.053). The SCAD vessels and segments demonstrated the expected vessel tapering over the length of the vessel, and there was no step-down in diameter at the SCAD-segment.

## Discussion

### Why does SCAD affect some arteries and not others?

It is not well understood why an individual with a genetically vulnerable coronary extracellular matrix,^4^ who experiences an environmental trigger, develops SCAD in a particular region or segment of their coronary artery tree.^1^

Our results support a multi-level susceptibility model for SCAD. At the whole-tree level, SCAD survivors showed greater curvature and higher TAESS and TSVI than controls, suggesting a global coronary geometry–flow phenotype rather than a purely focal abnormality. At the vessel (territory) level, SCAD-affected arteries also had smaller average diameters, along with higher curvature, TAESS, and TSVI, than matched control territories, consistent with the clinical observation that SCAD clusters in particular arteries and regions. Within individuals, lesion segments were characterized by smaller diameter and higher TAESS and TSVI than non-affected segments from the same tree, whereas curvature was only borderline different, indicating that curvature may be more informative at global/regional scales than as a lesion-local marker, and that shear metrics better capture lesion-level vulnerability. Only a minority of participants underwent CTCA during the acute event; however, we observed no meaningful differences between acute and non-acute CTCA subgroups in this cohort (**Supplement 6 Table S1**).

Our findings support a hypothetical model in which genetically mediated extracellular matrix fragility may interact with local hemodynamic stresses to precipitate vessel wall injury. Smaller-diameter arteries are typically associated with higher endothelial shear stress, which may provide an anatomical basis for the endothelial shear stress findings we observed. Chronically elevated and variable shear stress may induce maladaptive remodeling, weakening the arterial media and increasing susceptibility to intramural hemorrhage. Acute triggers that further spike endothelial shear stress may then precipitate vasa vasorum rupture resulting in clinical SCAD.

### Pathophysiology of high endothelial shear stress

High endothelial shear stress may play a role in SCAD pathophysiology, but further work is needed to ascertain if the high endothelial shear stress we have found predated the SCAD event. Endothelial shear stress plays a critical role in the development of the coronary arteries. A mechanosensitive ion channel, PIEZO2, specific to the coronary endothelium, guides the development of the coronary vasculature.^22^ There appears to be an optimal level of endothelial shear stress for coronary artery and cardiac function.

Chronic high endothelial shear stress can influence the remodeling of coronary vessels by affecting endothelial and vascular smooth muscle cells.^6^ Both these cell types are thought to play a role in the pathophysiology of SCAD. Endothelial cells are subjected to changes in blood flow and sense endothelial shear stress, and via the release of nitric oxide, regulate vascular smooth muscle tone and arterial resistance.^5^ In addition, high endothelial shear stress changes the gene expression profile of *ex vivo* endothelial cells. Relevant to SCAD, genes involved in matrix remodeling and angiogenesis are upregulated.^6,23^

High endothelial shear stress has been directly linked to arterial disease states. Higher segmental endothelial shear stress at sites of coronary plaque may increase plaque vulnerability,^24,25^ and similar mechanisms have been implicated in aneurysm formation.^6^ At an arterial segment level, chronically high shear stress is likely to produce a localized change in vessel wall architecture, resulting in adaptive extracellular matrix changes and subtle vessel remodeling, making that coronary artery segment more vulnerable to SCAD.

The inability to respond effectively to acute spikes in shear stress may be due to endothelial cells’ reduced responsiveness, with chronic nitric oxide upregulation in the high-shear-stress state.^6^

### Clinical applications

Our preliminary study may be the initial first step in extending a well-established paradigm of hemodynamic modelling (e.g., CT-derived fractional flow reserve) for future clinical decision-making to identify SCAD patients by means of a distinctive hemodynamic pattern, i.e., elevated TAESS and TSVI, coupled with adverse anatomy. Prior to clinical application, prospective validation against a clinical reference standard by integrating TAESS/TSVI via additional computational CTCA assessment is, therefore, an intuitive next step. Analogous to the emerging awareness that stenosis-only assessment for physiologic decision-making remains suboptimal, it can be hypothesized that operationalizing segment-level vulnerability detection in a setting where anatomy alone is insufficient, will be problematic. In this study, we begin to provide a critical proof-of-concept that establishes a clinically plausible pathway for SCAD risk stratification using anatomical and hemodynamic features, which is consistent with current post-SCAD management principles, including blood pressure control and avoidance of activities known to cause shear stress spikes. We are not suggesting that the current findings indicate that changes to these clinical factors will result in changes to the CFD parameters measured, but merely that the findings mechanistically support what is currently known about SCAD risk-reduction. As such, this model motivates a prospective study of whether hemodynamic surrogates relate to recurrence risk, with the potential to identify SCAD-affected vessels via non-invasive CTCA diagnostic, in a condition that readily lends itself to non-invasive modalities, given the high risk of iatrogenic dissection with invasive coronary angiography in patients who have suffered a SCAD, and recommendation to treat patients conservatively where possible. These proof-of-concept examples position computational hemodynamics as a promising next frontier in SCAD diagnostics.

### SCAD subtypes represent the same underlying vessel vulnerabilities

We found no difference between type 1 SCAD, which involves endothelial disruption (angiographic double lumen), and other types of SCAD, which consist solely of an intramural hematoma, across any of the parameters compared, including TAESS, suggesting they lie on the same mechanistic continuum. It has previously been demonstrated in a clinical coronary angiography follow-up study that SCAD cases due to an isolated intramural hematoma (type 2 SCAD) are at greater risk of progression, with 20% subsequently developing intimal dissection (type 1 SCAD) within 14 days.^26^ Therefore, it is probable that the underlying extracellular matrix alterations in the coronary artery wall render it susceptible to an environmental insult, such as increased shear stress, and thus, the development of SCAD, are the same in type 1 versus types 2 and 3 SCAD.

## Conclusions

This case-control study supports the notion that SCAD survivors have a unique coronary anatomy compared to sex- and ethnicity-matched controls. There are global differences at both the coronary tree and coronary territory levels compared to controls. Additionally, compared to the remaining coronary tree within an individual, SCAD segments had a characteristic profile. Coronary artery endothelial shear stress may play a critical role in the pathophysiology of all types of SCAD. Further prospective studies are required to validate these findings, prior to any major clinical applications.

## Limitations

This study involves the largest number of SCAD survivors with angiographically verified lesions, in which SCAD locations are carefully matched to segmented three-dimensional models derived from CTCA data. The study provides a complete CFD assessment, which was compared to a large, complete data set of controls. The results are potentially broadly generalizable to the typical SCAD patient. Despite these strengths, we acknowledge several limitations to our study. First, there is a significant age difference between SCAD survivors and controls, with the median age of controls being 10 years older than that of SCAD survivors. However, the direction of this difference adds further weight to our results of higher endothelial shear stress in the SCAD survivors because, as arteries age, wall stiffness increases with a higher proportion of collagen to elastin – collagen being 100 times stiffer than elastin – and consequently higher vascular stiffness and greater endothelial shear stress.^27^ It would therefore be expected that our reported differences would be further exacerbated with age-matched controls. Second, CTCA was performed at various times relative to the initial acute SCAD presentation. While most cases (26 of 36) had their CTCA performed after the SCAD event and showed a fully healed vessel anatomy, a smaller proportion (10 of 36) had the CTCA performed around the time of the acute SCAD episode. To address this issue, we compared these two groups but found no differences in any outcomes. Third, remodeling of the artery segment likely occurs after the SCAD episode, which may affect vessel parameters. This is an unknown confounding factor. Finally, the relatively small sample size reflects the rarity of SCAD and precluded independent validation of the multivariable discrimination analyses. Consequently, these findings should be considered exploratory and hypothesis-generating until confirmed in larger multicenter cohorts.

## Data Availability

The data are not publicly available due to privacy restrictions. De-identified data are available from the corresponding author upon reasonable request.

## Funding and Acknowledgements

We acknowledge the SCAD survivors in VASC, without whom this work would not have been possible. In particular, we acknowledge Sarah Ford, President of SCAD Research Inc, the SCADaddles and all SCAD survivors who have generously contributed to and supported our work.

This work was supported in part by grants from the Cardiac Society of Australia and New Zealand, the National Health and Medical Research Council, Australia (APP1161200), NSW Health, and SCAD Research Inc. J.C.K. acknowledges research support from the US National Institutes of Health (R01HL148167), Leducq Foundation, New South Wales Health grant RG194194, the Bourne Foundation, Snow Medical, and Agilent. R.M.G. is supported by a National Health and Medical Research Council L3 Investigator Grant (APP2010203) and a New South Wales Health Cardiovascular Senior Scientist Grant. L.M-C. acknowledges funding support from a National Health and Medical Research Council, Australia, Postgraduate Scholarship (GNT2013809) with co-funding from a National Heart Foundation PhD Scholarship (106228), a Royal Australasian College of Physicians Research Establishment Fellowship (2026REF056) and UNSW FMH Collaborative Seed Grant.

## Abbreviations

ACS: acute coronary syndrome
CAD: coronary artery disease
CFD: computational fluid dynamics
CTCA: computed tomographic coronary angiography
ICA: invasive coronary angiogram
LAD: left anterior descending
LCx: left circumflex
OSI: oscillatory shear index
RCA: right coronary artery
ROC: receiver operating characteristic
RRT: relative residence time
SCAD: spontaneous coronary artery dissection
TAESS: time-averaged endothelial shear stress
TSVI: topological shear variation index

